# Diagnosis and treatment of opioid-related disorders in a South African private sector medical insurance scheme: a cohort study

**DOI:** 10.1101/2022.04.28.22274253

**Authors:** Mpho Tlali, Andrew Scheibe, Yann Ruffieux, Morna Cornell, Anja E Wettstein, Matthias Egger, Mary-Ann Davies, Gary Maartens, Leigh F Johnson, Andreas D Haas

## Abstract

**Background:** The use of opioids is increasing globally, but data from low- and middle-income countries on opioid-related mental and behavioural disorders (hereafter referred to as opioid-related disorders) are scarce. This study examines the incidence of opioid-related disorders, opioid agonist use, and excess mortality among persons with opioid-related disorders in South Africa’s private healthcare sector.

**Methods:** We analysed longitudinal data of beneficiaries (≥ 11 years) of a South African medical insurance scheme using reimbursement claims from Jan 1, 2011, to Jul 1, 2020. Beneficiaries were classified as having an opioid-related disorder if they received an opioid agonist (buprenorphine or methadone) or an ICD-10 diagnosis for harmful opioid use (F11.1), opioid dependence or withdrawal (F11.2-4), or an unspecified or other opioid-related disorder (F11.0, F11.5-9). We calculated adjusted hazard ratios (aHR) for factors associated with opioid-related disorders, estimated the cumulative incidence of opioid agonist use after receiving an ICD-10 diagnosis for opioid dependence or withdrawal, and examined excess mortality among beneficiaries with opioid-related disorders.

**Results:** Of 1,251,458 beneficiaries, 1,286 (0.1%) had opioid-related disorders. Between 2011 and 2020, the incidence of opioid-related disorders increased by 12% (95% CI 9%-15%) per year. Men, young adults in their twenties, and beneficiaries with co-morbid mental health or other substance use disorders were at increased risk of opioid-related disorders. The cumulative incidence of opioid agonist use among beneficiaries who received an ICD-10 diagnosis for opioid dependence or withdrawal was 18.0% (95% CI 14.0-22.4) 3 years after diagnosis. After adjusting for age, sex, year, medical insurance coverage, and population group, opioid-related disorders were associated with an increased risk of mortality (aHR 2.28, 95% CI 1.84-2.82). Opioid-related disorders were associated with a 7.8-year shorter life expectancy.

**Conclusions:** The incidence of patients diagnosed with or treated for an opioid-related disorder in the private sector is increasing rapidly. People with opioid-related disorders are a vulnerable population with substantial psychiatric comorbidity who often die prematurely. Evidence-based management of opioid-related disorders is urgently needed to improve the health outcomes of people with opioid-related disorders.

## Background

The use of opioids is increasing worldwide (UNODC, 2021). In 2019, the estimated global prevalence of non-medical opioid use was 1.2% in adults aged 15-64 years (United Nations Office on Drugs and Crime, 2021). The World Health Organization (WHO) classifies harmful opioid use, opioid dependence, withdrawal and other patterns or consequences of opioid use under the umbrella term opioid-related mental and behavioural disorders (hereafter referred to as opioid-related disorders) (WHO, 2016). Opioid dependence may develop after repeated or continuous use, with associated neurobiological, behavioural and psychological changes (WHO, 2009). Opioid dependence (in North America often referred to as opioid use disorder according to 5th edition of the American Psychiatric Association’s Diagnostic and Statistical Manual of Mental Disorders) (Degenhardt et al., 2019) is an important contributor to the global burden of disease, accounting for 70% of the disease burden from drug-related causes in 2019 (United Nations Office on Drugs and Crime, 2021). Mortality rates among people with an opioid dependence are higher than in the general population (Degenhardt et al., 2011). Globally, overdose and infectious disease complications (mainly due to HIV and viral hepatitis C infection) are the most important causes of mortality in people with an opioid-related disorders (Degenhardt et al., 2011). Opioid-related disorders commonly occur with other mental and substance use disorders, adding to potential harm and challenges in management (Abbafati et al., 2020a).

The WHO recommends opioid agonist maintenance therapy (OAMT) for the treatment of opioid dependence (WHO, 2009). Methadone and buprenorphine are the most widely used agonist medications for OAMT. There is strong evidence from randomised controlled trials that OAMT is more effective in retaining participants in treatment and suppressing opioid use than nonDpharmacological approaches (Mattick et al., 2009, 2014). A Cochrane review comparing methadone maintenance therapy versus no opioid replacement therapy for opioid dependence shows that methadone maintenance therapy reduces the risk of morphine positive urine or hair analysis at follow-up by 34% (relative risk 0.66, 95% CI 0.56, 0.78) (Mattick et al., 2009). A further Cochrane review showed that buprenorphine maintenance treatment of heroin dependence is superior in suppressing illicit opioid use at high doses of 16 mg or greater compared to placebo (Mattick et al., 2014).

### South African context

#### Private medical insurance

South Africa has a dual health care system with large disparities between the public and private sectors (Day & Gray, 2013; Hassim et al., 2007). In 2018, 15% of the population was covered by private sector medical insurance (Day & Gray, 2013). However, a significant fraction of the population accesses private healthcare without medical insurance and similarly, many people with private medical insurance access both private and public healthcare (Hassim et al., 2007). People with medical insurance generally have higher socio-economic status than people who access public healthcare services (Hassim et al., 2007).

The South African Council for Medical Schemes defines the benefits that private medical insurance schemes need to provide to all scheme members. These prescribed minimum benefits (PMBs) cover the cost of care (medical services and medications) for emergencies and selected acute and chronic medical conditions (Council for Medical Schemes, 2020). Beneficiaries of medical insurance schemes can pay the cost of medications and medical management for conditions not included in the PMB from available benefits. Benefits are fixed annual amounts to cover for consultations with general practitioners, specialist, acute medicine, laboratory tests and other medical expenses. The benefits of medical insurance packages increase with higher premiums (Council for Medical Schemes, 2020). If benefits are exhausted, beneficiaries have to cover medical costs from the own resources.

#### Epidemiology of opioid use

Data on opioid use in South Africa are limited. In 2012, 0.3% of people aged ≥15 years were estimated to have used heroin, the most widely used opioid (known locally as nyaope, whoonga, unga, or sugars) in the previous three months, according to a national household survey (Peltzer & Phaswana-Mafuya, 2018). Past year use of opioids was estimated at 0.5% (L Weich et al., 2017). Programmatic and research data suggest opioid use has increased across the country in the past decade (Dada et al., 2019; Harker et al., 2020; Peltzer et al., 2010). The increased affordability and accessibility of heroin accounts for the surge in opioid use locally and across sub-Saharan Africa (Eligh, 2010; Mokwena, 2016). Over-the-counter (OTC) prescription opioids, such as codeine and tramadol, are also used in many sub-Saharan African countries for non-medical purposes (Parry et al., 2017; United Nations Office on Drugs and Crime, 2021). Non-medical use of codeine in South Africa is reported in 2% of adults, but account for a minority of people with opioid dependence (National Department of Health (NDoH), Statistictics South Africa (Stats SA), South African Medical Research Council (SAMRC), 2018).

#### Management of opioid dependence

Few people with opioid dependence in South Africa receive evidence-based interventions (Scheibe, Shelly, & Versfeld, 2020). By 2020, only one methadone, one buprenorphine and one buprenorphine-naloxone product were registered for OAMT (L Weich et al., 2017). The cost of methadone is 10 – 30 times higher than in other middle-income counties (Scheibe et al., 2018). Methadone and buprenorphine are not included in the South African National Department of Health’s Essential Medicines List. Therefore, these drugs are not available in the public healthcare sector (Scheibe et al., 2018). In the private health care sector, methadone and buprenorphine are generally available, but the coverage of the high cost of these medications by medical insurance schemes is limited. Medications used during psychiatric in-hospital stays for ‘abuse or dependence of psychoactive drugs’ are covered for a maximum of three weeks per year as a PMB. Medications used during outpatient care have to be covered by beneficiaries’ annual benefits (Council for Medical Schemes, 2020). Depending on the plan, annual benefits for out-of-hospital care could cover approximately 3 months of methadone at a recommended dose (OpenUp, 2022). The limited capacity and awareness among medical practitioners on the use of OAMT is an additional factor limiting access (Scheibe et al., 2018).

Despite their ineffectiveness, most government and non-governmental drug treatment centres provide detoxification and abstinence-based rehabilitation to manage opioid dependence (Dada et al., 2021). For example, only 11.6% of opioid users (n=534) were retained for two months in a public-funded, abstinence-based outpatient programme in Cape Town (Magidson et al., 2017). In another study in Gauteng province, 70.7% of clients (n=300) returned to daily heroin use within three months of discharge from a government in-patient detoxification centre (Morgan et al., 2019).

The largest number of OAMT clients receive treatment as part of harm reduction programmes provided by civil society organisations and academic institutions across five cities and funded by donors and one metropolitan municipality (Dada et al., 2021). These harm reduction programmes target socio-economically disadvantaged people (particularly those living on the street). At the end of June 2020, 885 people were on OAMT in community-based harm reduction programmes (Dada et al., 2021). To our knowledge, there are no published reports of substance use disorder treatment in South Africa’s private (for-profit) sector.

This study examined the incidence of diagnosed harmful opioid use, opioid dependence or withdrawal, use of opioid agonists, and mortality of persons with opioid-related disorders among beneficiaries of a private sector medical insurance scheme in South Africa.

## Methods

### Study design

We conducted a cohort study using outpatient, hospital, and medication reimbursement claims of beneficiaries of a South African medical insurance scheme, and data of the vital status of beneficiaries from the National Population Register (NPR).

### Data sources

We analysed outpatient, hospital, and medication claim data of beneficiaries of a South African medical insurance scheme, covering the period from Jan 1, 2011, to Jul 1, 2020. Outpatient and hospital claims include International Classification of Diseases, 10th Revision (ICD-10) diagnoses (ICD-11 codes only became effective on Jan 1, 2022 and were not in use throughout the duration of this analysis). Medication claims include medication names, classifications (Anatomical Therapeutic Chemical [ATC] code), strength, the dispensed amount, and the date of the claim but no information on the dosing. The medical scheme administrator obtained data on the vital status of beneficiaries from the NPR covering the period from Jan 1, 2011, to Jan 26, 2021. Data on the vital status of beneficiaries collected previously by the medical insurance scheme were updated with the NPR data.

### Study participants

Individuals aged 11 years or older who had health care coverage with the medical insurance scheme were eligible for analysis. Persons without health insurance who are accessing public sector health care are not included in our study. Beneficiaries with unknown sex or age were excluded (Figure S1). We restricted the analysis of the cumulative incidence of opioid agonist therapy to beneficiaries who had received an ICD-10 diagnosis of opioid dependence or withdrawal (F11.2-4) (Figure S1). Beneficiaries who could not be linked to NPR data were excluded from mortality analyses (Figure S1).

### Measures and procedures

We defined baseline as beneficiaries’ date of enrolment with the medical insurance scheme, Jan 1, 2011, or their 11^th^ birthday, whichever occurred last. We classified beneficiaries as having an opioid-related disorder if they had received an ICD-10 F11 diagnosis for (1) harmful opioid use (F11.1), (2) opioid dependence or withdrawal (F11.2-4), (3) or an unspecific or other opioid-related disorder (F11.0, F11.5-9) or (4) were prescribed one of the following opioid agonists: buprenorphine sub-lingual tablet (2mg or 8 mg, ATC code N07BC01), methadone solution (2mg/1ml, ATC code N07BC02) or buprenorphine-naloxone (2mg/0.5mg or 8mg/2mg, ATC code N07BC51) sublingual tablets. Beneficiaries who received methadone suspension (2mg/5ml) only (ATC code N07BC02) were not classified as having opioid-related disorders because this formulation is often used for other indications (L Weich et al., 2017). In our analysis, patients classified as having an opioid-related disorder do not necessarily meet the Diagnostic and Statistical Manual of Mental Disorders, Fifth Edition (DSM-5) criteria for OUD. The cause of death was classified as unnatural (ICD-10 codes V01–Y98) or natural as per the ICD-10. Opioid overdose (ICD-10 X42 or X62) is coded as an unnatural cause of death.

Substance use disorders (ICD-10 codes F10-F19) were classified as alcohol use disorder (ICD-10 code F10), opioid-related disorders (ICD-10 code F11), multiple drug use disorder (ICD-10 code F19), or other substance use disorders (ICD-10 codes F12-F18). Mental health disorders (ICD-10 codes F00-F09, F20-F99) were classified as serious mental disorders (ICD-10 codes F20-F29, F31), depression (ICD-10 codes F32, F33, F34.1), anxiety disorders (ICD-10 codes F40-48), or other mental disorders (ICD-10 codes F00-F09, F50-F99). We grouped age in 10 years age brackets (11-19, 20-29, 30-39, 40-49, 50-59, 60-69, and 70+) and year in 3-year brackets (2011-2013, 2014-2016, 2017-2019, and 2020-2021). Infectious diseases and infections were grouped into HIV (ICD-10 codes B20-24), hepatitis C virus (ICD-10 codes B17.1, B18.2), tuberculosis (ICD-10 codes A15-A19) and infective endocarditis (ICD-10 code I33.0).

### Statistical analysis

We described the characteristics of the study population using summary statistics. Using Cox proportional hazards models, we estimated unadjusted and adjusted hazard ratios (HR) and 95% confidence intervals (CI) for factors associated with OUD, F11 diagnoses, and opioid agonist use. In this analysis, we followed beneficiaries from baseline to the end of their health care plan or the outcome of interest (whichever occurred first). Variables considered in univariable and multivariable analyses were chosen a priori: year, sex, age group, population group, serious mental disorders, depression, anxiety, other mental disorders, alcohol use disorders, and other substance use disorders. We modelled age group, year (as a continuous variable), and mental and substance use diagnoses as time-varying covariates. We estimated the cumulative incidence of opioid agonist use (ATC code N07BC) after F11 diagnosis considering mortality as a competing event (Coviello & Boggess, 2004; Gooley et al., 1999). We followed beneficiaries from opioid use disorder diagnosis to opioid agonist use, death, or end of the follow-up period. We estimated unadjusted and adjusted HRs and 95% CIs for factors associated with mortality using Cox proportional hazards models. We modelled age, year, and substance use diagnoses as time-varying covariates. We modelled year as a categorical predictor to adjust for excess mortality in 2020-2021 due to the COVID-19 epidemic. In the analysis of factors associated with mortality, we followed beneficiaries from baseline to database closure (Jan 26, 2021) or death (whichever occurred first). The proportional hazards assumption was assessed based on Schoenfeld residuals and visual inspection of log-log plots of survival. Finally, we estimated the excess life-years lost associated with OUD before the age of 85. The excess life-years lost measures how many life-years someone with a given exposure is expected to lose starting from the age of onset of the exposure compared to someone of the same age who is unexposed (Andersen, 2017; Plana-Ripoll et al., 2019). We calculated excess life-years lost stratified by sex and overall. We disaggregated the excess LYL into natural and unnatural death components. We used bootstrap simulation to produce 95% confidence intervals. Calculations of the excess life-years lost were performed using the R package *lillies* (Plana-Ripoll et al., 2020).

### Ethical considerations

We obtained data from the International Epidemiology Database to Evaluate AIDS (IeDEA) Southern Africa collaboration (Chammartin et al., 2020). IeDEA Southern Africa collects routine clinical and administrative data for HIV-positive and HIV-negative children and adults in six Southern African countries. All programmes participating in IeDEA have ethics approval to contribute de-identified data to the IeDEA Data Centre. Beneficiaries of the medical insurance scheme or their guardians provided consent for their data to be used in research. The Human Research Ethics Committee of the University of Cape Town, South Africa (reference number 084/2006), and the Cantonal Ethics Committee of the Canton of Bern, Switzerland (150/14, PB_2016-00273) granted permission to analyse data.

## Results

Our database included 1,549,123 persons. We excluded 279,430 children younger than 11 years (18.0%) and 18,235 persons (1.2%) of unknown age or sex (Figure S1). The remaining 1,251,458 beneficiaries were eligible for this analysis, among whom 1,286 (0.1%) had an opioid-related disorder. The median follow-up time was 3.2 years (IQR 1.2-6.3). The median age of the beneficiaries at baseline was 33 years (IQR 21-47), 51.9% were women, 24.9% had received a diagnosis of a mental health disorder, 0.6% had a substance use disorder related to substances other than opioids, and 6.5% had an HIV diagnosis. Beneficiaries with opioid-related disorders were of similar age (median 32 years, IQR 21-47), more likely to be male (63.0%), had a higher prevalence of mental health diagnosis (69.5%), additional substance use disorder diagnoses (related to substances other than opioids) (31.4%), and infectious diseases diagnosis (11.7%) (HIV (9.5%), Hepatitis C (HCV) (0.7%), and TB (3.4%)) than those without opioid-related disorders. Few persons with and without opioid-related disorders had been diagnosed with infective endocarditis (Table 1).

**Table 1:** Characteristics of beneficiaries of a South African medical insurance scheme with and without opioid-related disorder.

|  | Persons with<br>opioid-related<br>disorder |  | Persons without<br>opioid-related disorder |  | Total |  |
| --- | --- | --- | --- | --- | --- | --- |
|  | N=1,286 | (0.1) | N=1,250,172 | (99.9) | N=1,251,458 | (100.0) |
| Sex |  |  |  |  |  |  |
| Male | 810 | (63.0) | 601,610 | (48.1) | 602,420 | (48.1) |
| Female | 476 | (37.0) | 648,562 | (51.9) | 649,038 | (51.9) |
| Age at baseline, years |  |  |  |  |  |  |
| 11-19 | 287 | (22.3) | 289,441 | (23.2) | 289,728 | (23.2) |
| 20-29 | 279 | (21.7) | 240,103 | (19.2) | 240,382 | (19.2) |
| 30-39 | 252 | (19.6) | 256,266 | (20.5) | 256,518 | (20.5) |
| 40-49 | 187 | (14.5) | 202,471 | (16.2) | 202,658 | (16.2) |
| 50-59 | 159 | (12.4) | 149,516 | (12.0) | 149,675 | (12.0) |
| 60+ | 122 | (9.5) | 112,375 | (9.0) | 112,497 | (9.0) |
| Median (IRQ) | 32 | (21-47) | 33 | (21-47) | 33 | (21-47) |
| Population group |  |  |  |  |  |  |
| Indian | 71 | (5.5) | 54,399 | (4.4) | 54,470 | (4.4) |
| Black | 568 | (44.2) | 662,933 | (53.0) | 663,501 | (53.0) |
| Mixed | 104 | (8.1) | 80,111 | (6.4) | 80,215 | (6.4) |
| White | 298 | (23.2) | 217,277 | (17.4) | 217,575 | (17.4) |
| Missing | 245 | (19.1) | 235,452 | (18.8) | 235,697 | (18.8) |
| Mental health diagnosis | 894 | (69.5) | 311,003 | (24.9) | 311,897 | (24.9) |
| Serious mental disorder | 278 | (21.6) | 23,773 | (1.9) | 24,051 | (1.9) |
| Depression | 680 | (52.9) | 164,396 | (13.1) | 165,076 | (13.2) |
| Anxiety disorder | 568 | (44.2) | 195,045 | (15.6) | 195,613 | (15.6) |
| Other mental disorder | 277 | (21.5) | 70,142 | (5.6) | 70,419 | (5.6) |
| Substance use disorder diagnosis (excl. ICD-10 F11) | 404 | (31.4) | 7,456 | (0.6) | 7,860 | (0.6) |
| Alcohol use disorder (ICD-10 F10) | 90 | (7.0) | 3,911 | (0.3) | 4,001 | (0.3) |
| Multiple drug use (ICD-10 F19) | 301 | (23.4) | 2,478 | (0.2) | 2,779 | (0.2) |
| Other substance use disorders (ICD-10 F12-F18) | 163 | (12.7) | 2,090 | (0.2) | 2,253 | (0.2) |
| Infectious diseases | 151 | (11.7) | 89,429 | (7.2) | 89,580 | (7.2) |
| HIV | 122 | (9.5) | 81,266 | (6.5) | 81,388 | (6.5) |
| HCV | 9 | (0.7) | 121 | (0.0) | 130 | (0.0) |
| TB | 44 | (3.4) | 17,147 | (1.4) | 17,191 | (1.4) |
| Infective endocarditis | 3 | (0.2) | 349 | (0.0) | 352 | (0.0) |
| Proxies for opioid-related disorder |  |  |  |  |  |  |
| Harmful use | 122 | (9.5) | 0 | (0.0) | 122 | (0.0) |
| Dependence or withdrawal | 458 | (35.6) | 0 | (0.0) | 458 | (0.0) |
| Unspecified or other F11 diagnosis | 287 | (22.3) | 0 | (0.0) | 287 | (0.0) |
| Opioid agonist use | 526 | (40.9) | 0 | (0.0) | 526 | (0.0) |
| Methadone | 523 | (40.7) | 0 | (0.0) | 523 | (0.0) |
| Buprenorphine | 4 | (0.3) | 0 | (0.0) | 4 | (0.0) |
| Buprenorphine-Naloxone | 0 | (0.0) | 0 | (0.0) | 0 | (0.0) |

Of the 1,286 persons classified as having an opioid-related disorder, 122 (9.5%) received an ICD-10 diagnosis for harmful opioid use, 458 (35.6%) for opioid dependence or withdrawal, 287 (22.3%) and 526 (40.9%) had received an opioid agonist. Methadone and buprenorphine were prescribed to 523 (40.7%) and 4 (0.3%) beneficiaries. Buprenorphine in combination with naloxone was not prescribed in this cohort (Table 1). The median total amount of methadone dispensed per beneficiary during the entire study duration was 400 mg (IQR 120-1080).

Unadjusted and adjusted HRs for factors associated with opioid-related disorders, harmful opioid use, opioid dependence or withdrawal, unspecified or other F11 diagnoses, and opioid agonist use are shown in Table 2. From 2011 to 2020, the incidence of opioid-related disorders increased by 12% per year (aHR 1.12 [95% CI 1.09-1.15]), harmful opioid use diagnoses by 16% per year (aHR 1.16 [95% CI 1.07-1.26]), opioid dependence and withdrawal diagnoses by 8% per year (aHR 1.08 [95% CI 1.04-1.13]) and opioid agonist use by 17% per year (aHR 1.17 [95% CI 1.12-1.22]). Men and adults 20-39 years had a higher incidence of opioid-related disorders, opioid dependence and withdrawal, and opioid agonist use compared to women and adults 40-49 years. Persons with depression, anxiety disorders, serious mental disorders, and other substance use disorders (F12-F18) had a higher risk of harmful opioid use, opioid dependence or withdrawal, unspecified or other F11 diagnoses, and opioid agonist compared to persons without these conditions (Table 2).

**Table 2.**
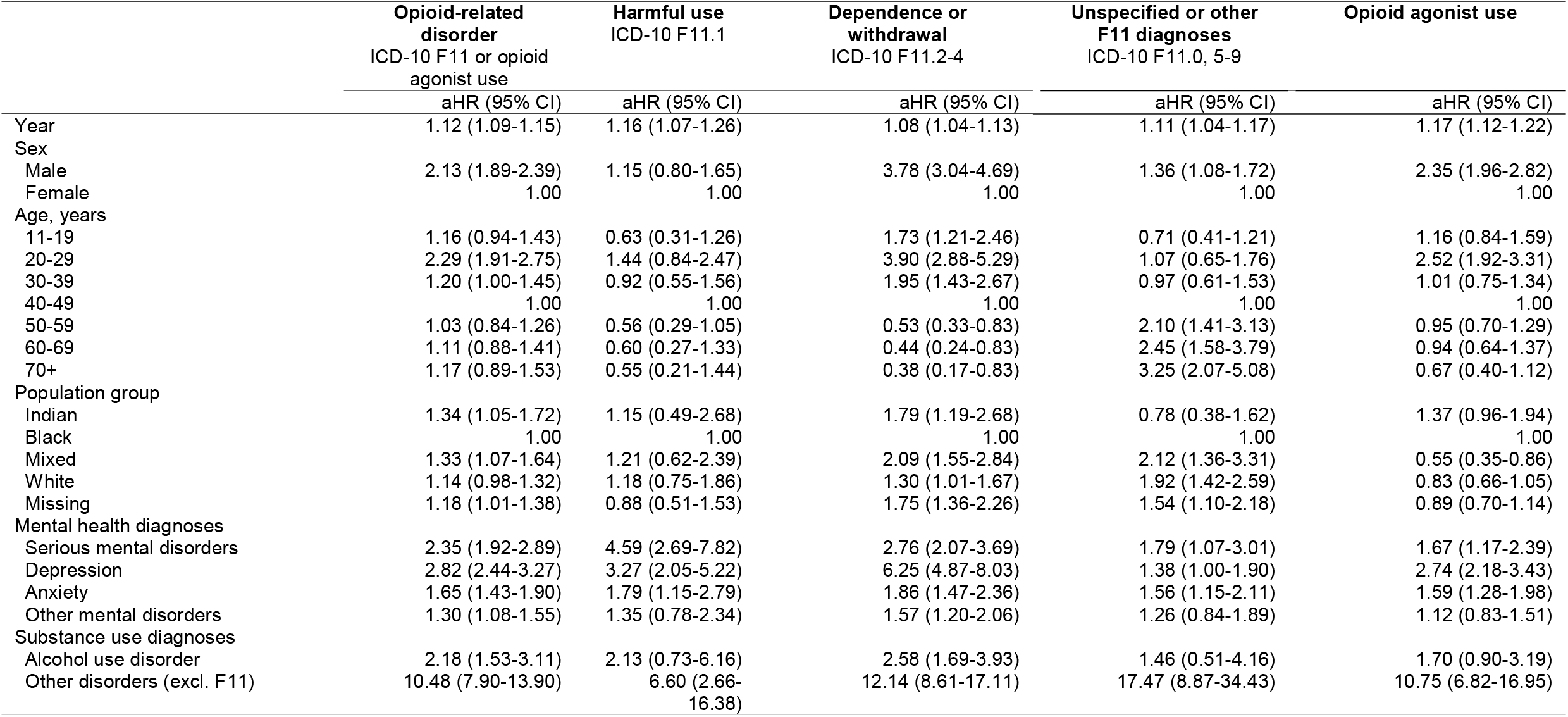
Hazard ratios for factors associated with opioid-related disorder among beneficiaries of a South African medical insurance scheme.

|  | Opioid-related disorder<br>ICD-10 F11 or opioid<br>agonist use | Harmful use<br>ICD-10 F11.1 | Dependence or<br>withdrawal<br>ICD-10 F11.2-4 | Unspecified or other<br>F11 diagnoses<br>ICD-10 F11.0, 5-9 | Opioid agonist use |
| --- | --- | --- | --- | --- | --- |
|  | aHR (95% CI) | aHR (95% CI) | aHR (95% CI) | aHR (95% CI) | aHR (95% CI) |
| Year | 1.12 (1.09-1.15) | 1.16 (1.07-1.26) | 1.08 (1.04-1.13) | 1.11 (1.04-1.17) | 1.17 (1.12-1.22) |
| Sex |  |  |  |  |  |
| Male | 2.13 (1.89-2.39) | 1.15 (0.80-1.65) | 3.78 (3.04-4.69) | 1.36 (1.08-1.72) | 2.35 (1.96-2.82) |
| Female | 1.00 | 1.00 | 1.00 | 1.00 | 1.00 |
| Age, years |  |  |  |  |  |
| 11-19 | 1.16 (0.94-1.43) | 0.63 (0.31-1.26) | 1.73 (1.21-2.46) | 0.71 (0.41-1.21) | 1.16 (0.84-1.59) |
| 20-29 | 2.29 (1.91-2.75) | 1.44 (0.84-2.47) | 3.90 (2.88-5.29) | 1.07 (0.65-1.76) | 2.52 (1.92-3.31) |
| 30-39 | 1.20 (1.00-1.45) | 0.92 (0.55-1.56) | 1.95 (1.43-2.67) | 0.97 (0.61-1.53) | 1.01 (0.75-1.34) |
| 40-49 | 1.00 | 1.00 | 1.00 | 1.00 | 1.00 |
| 50-59 | 1.03 (0.84-1.26) | 0.56 (0.29-1.05) | 0.53 (0.33-0.83) | 2.10 (1.41-3.13) | 0.95 (0.70-1.29) |
| 60-69 | 1.11 (0.88-1.41) | 0.60 (0.27-1.33) | 0.44 (0.24-0.83) | 2.45 (1.58-3.79) | 0.94 (0.64-1.37) |
| 70+ | 1.17 (0.89-1.53) | 0.55 (0.21-1.44) | 0.38 (0.17-0.83) | 3.25 (2.07-5.08) | 0.67 (0.40-1.12) |
| Population group |  |  |  |  |  |
| Indian | 1.34 (1.05-1.72) | 1.15 (0.49-2.68) | 1.79 (1.19-2.68) | 0.78 (0.38-1.62) | 1.37 (0.96-1.94) |
| Black | 1.00 | 1.00 | 1.00 | 1.00 | 1.00 |
| Mixed | 1.33 (1.07-1.64) | 1.21 (0.62-2.39) | 2.09 (1.55-2.84) | 2.12 (1.36-3.31) | 0.55 (0.35-0.86) |
| White | 1.14 (0.98-1.32) | 1.18 (0.75-1.86) | 1.30 (1.01-1.67) | 1.92 (1.42-2.59) | 0.83 (0.66-1.05) |
| Missing | 1.18 (1.01-1.38) | 0.88 (0.51-1.53) | 1.75 (1.36-2.26) | 1.54 (1.10-2.18) | 0.89 (0.70-1.14) |
| Mental health diagnoses |  |  |  |  |  |
| Serious mental disorders | 2.35 (1.92-2.89) | 4.59 (2.69-7.82) | 2.76 (2.07-3.69) | 1.79 (1.07-3.01) | 1.67 (1.17-2.39) |
| Depression | 2.82 (2.44-3.27) | 3.27 (2.05-5.22) | 6.25 (4.87-8.03) | 1.38 (1.00-1.90) | 2.74 (2.18-3.43) |
| Anxiety | 1.65 (1.43-1.90) | 1.79 (1.15-2.79) | 1.86 (1.47-2.36) | 1.56 (1.15-2.11) | 1.59 (1.28-1.98) |
| Other mental disorders | 1.30 (1.08-1.55) | 1.35 (0.78-2.34) | 1.57 (1.20-2.06) | 1.26 (0.84-1.89) | 1.12 (0.83-1.51) |
| Substance use diagnoses |  |  |  |  |  |
| Alcohol use disorder | 2.18 (1.53-3.11) | 2.13 (0.73-6.16) | 2.58 (1.69-3.93) | 1.46 (0.51-4.16) | 1.70 (0.90-3.19) |
| Other disorders (excl. F11) | 10.48 (7.90-13.90) | 6.60 (2.66-16.38) | 12.14 (8.61-17.11) | 17.47 (8.87-34.43) | 10.75 (6.82-16.95) |

The cumulative incidence of opioid agonist use among beneficiaries who received an ICD-10 diagnosis for opioid dependence or withdrawal was 18.0% (95% CI 14.0-22.4) 3 years after diagnosis. (Table 3). The cumulative incidence of opioid agonist use was higher in men (20.2% [95% CI 15.4-25.4]) than in women (11.2% (95% CI 5.0-20.0)]) (Table 3).

**Table 3.** Cumulative incidence of opioid agonist use among beneficiaries who received an ICD-10 diagnosis for opioid dependence or withdrawal.

| Years after diagnosis | Opioid agonist use<br>Cumulative incidence, % (95% CI) |  |  |
| --- | --- | --- | --- |
|  | Men | Women | Total |
| 0 | 10.5 (7.5-14.0) | 6.1 (2.7-11.6) | 9.4 (6.9-12.3) |
| 1 | 15.8 (12.0-20.0) | 8.2 (4.0-14.4) | 13.9 (10.8-17.4) |
| 2 | 18.5 (14.2-23.3) | 8.2 (4.0-14.4) | 16.0 (12.5-20.0) |
| 3 | 20.2 (15.4-25.4) | 11.2 (5.0-20.0) | 18.0 (14.0-22.4) |

Analysis includes 458 beneficiaries who received in International Classification of Diseases, 10th Revision (ICD-10) diagnoses for opioid dependence (F11.2) or withdrawal (F11.3-4).

Of the 1,251,458 beneficiaries included in the study, 1,166,636 (93.2%) could be linked to the NPR and were included in the analysis of factors associated with mortality and excess mortality among persons with opioid-related disorders (Figure S1). Table 4 shows unadjusted and adjusted hazard ratios for factors associated with mortality. Beneficiaries with opioid-related disorders had a substantially increased risk of mortality (aHR 2.28 [95% CI 1.84-2.82]) after adjusting for year, age, sex, current medical insurance coverage, and population group (<u>model 1</u>). When also adjusting for co-morbid mental health and substance use disorder, opioid-related disorders remained associated with mortality (aHR 1.67 [95% CI 1.35-2.06]) (<u>model 2</u>), but the strength of the association was attenuated. Out of the four proxies used to identify persons with opioid-related disorders, opioid agonist use was the strongest risk factor for mortality (aHR 3.06, 95% CI 2.26-4.15), followed by harmful opioid use diagnosis (aHR 2.45, 95% CI 1.27-4.74), opioid dependence or withdrawal diagnosis (aHR 2.44, 95% CI 1.60-3.27) (<u>model 3</u>).

**Table 4.** Hazard ratios for mortality in persons with opioid-related disorders.

|  | Unadjusted analyses | Adjusted analyses |  |  |
| --- | --- | --- | --- | --- |
|  |  | Model 1 | Model 2 | Model 3 |
|  | HR<br>(95% CI) | aHR<br>(95% CI) | aHR<br>(95% CI) | aHR<br>(95% CI) |
| Opioid-related disorders | 2.13 (1.72-2.63) | 2.28 (1.84-2.82) | 1.67 (1.35-2.06) |  |
| Harmful use | 2.29 (1.19-4.39) |  |  | 2.45 (1.27-4.74) |
| Dependence or withdrawal | 1.60 (1.06-2.41) |  |  | 2.44 (1.60-3.72) |
| Unspecified or other F11 diagnosis | 2.38 (1.58-3.59) |  |  | 1.35 (0.89-2.03) |
| Opioid agonist use | 2.81 (2.09-3.77) |  |  | 3.06 (2.26-4.15) |

Model 1 adjusted for year, age, sex, medical insurance coverage, and population group. Model 2 adjusted for year, age, sex, medical insurance coverage, population group and co-morbid mental health and substance use disorder. Model 3 included binary exposure variables for ICD-10 diagnoses for harmful opioid use (F11.1), dependence or withdrawal (F11.2-4), unspecified or other opioid-related disorders (F11.0, F11.5-9) and a binary exposure variable for opioid agonist use and was adjusted for year, age, sex, medical insurance coverage, and population group.

Table 5 shows the excess life-years lost associated with opioid-related disorders. On average, persons with opioid-related disorders lost 7.8 life years before the age of 85 compared to persons without opioid-related disorders (men: 7.81 life years [95% CI 3.20-12.25], and women 7.83 life years [95% CI 3.14-11.90]). These excess life-years lost were almost entirely driven by natural deaths.

**Table 5.** Excess life-years lost associated with opioid-related disorders for natural, unnatural and all causes of death.

| **Sex** | **Excess life-years lost (95% confidence intervals)** |  |  |
| --- | --- | --- | --- |
|  | All-cause | Natural deaths | Unnatural deaths |
| Men | 7.81 (3.20-12.25) | 8.38 (3.84-12.72) | -0.57 (-1.54-1.56) |
| Women | 7.83 (3.14-11.90) | 7.52 (2.64-11.90) | 0.31 (-0.33-1.62) |

## Discussion

The findings from our analysis point to a notable increase in opioid use in South Africa. We found high yearly increases in the rate of opioid-related disorders, with a doubling in incidence from 2011 to 2020. Our analysis also highlights the major gaps in the management of opioid dependence in South Africa’s private healthcare sector. Males and young adults were identified as important risk groups for opioid-related disorders, bearing a disproportionate disease burden. A strong association was found between opioid-related disorders and psychiatric comorbidities, particularly for serious mental health disorders, depression, and anxiety disorders. Less than 20% of beneficiaries were prescribed opioid agonists within three years after receiving an ICD-10 diagnosis for opioid dependence or withdrawal. The total median amount of prescribed methadone suggests that most people only received agonist medications for a short period of time, clinically equivalent to withdrawal management rather than OAMT. Furthermore, we observed substantial excess mortality in persons with opioid-related disorders compared to those without opioid-related disorders.

The doubling of opioid-related disorders over the last decade observed in this study cohort is consistent with other global trends (Abbafati et al., 2020b; United Nations Office on Drugs and Crime, 2021). The United Nations Office on Drugs and Crime (UNODC) reported a near doubling of opioid use globally, driven by increased use in Asia and Africa in recent years (United Nations Office on Drugs and Crime, 2021).

Men and young adults aged 20-29 have been identified as important risk groups for opioid-related disorders. In South Africa, the average age of individuals accessing treatment for opioid dependence at abstinence-based drug treatment centres is around 30 years (Dada et al., 2019; Harker et al., 2020; Peltzer & Phaswana-Mafuya, 2018). Males comprise more than three-quarters of individuals treated for opioid dependence in South Africa (Dada et al., 2019; Harker et al., 2020; Ramlagan et al., 2010). Young men from socially disadvantaged communities are at risk of engaging in drug use for many reasons, including unemployment, limited social mobility, and low personal aspirations (Peltzer et al., 2010). The increased risk in men and young people for opioid dependence has also been reported in other global settings (Degenhardt et al., 2014, 2019).

Our results also show strong associations between opioid-related disorders, mental health, and other substance use disorders. The co-occurrence of opioid-related disorders and other mental and substance use disorders is well recognised (Jones & McCance-Katz, 2019; Kurth et al., 2018; L Dannatt, K J Cloete, M Kidd, 2014; Ramlagan et al., 2010). A South African study found that among people treated for opioid dependence in a public detoxification centre, 52% had concurrent use of methamphetamine, 26% had major depressive disorder, 20% had an anxiety disorder, and 8% had post-traumatic stress disorder (L Dannatt, K J Cloete, M Kidd, 2014). A recent US national survey showed that co-occurrence of mental health and substance use disorder is common among adults meeting DSM-IV criteria for opioid use disorder. Co-occurring substance use disorders ranged from 26.4% for alcohol use to 10.6% for methamphetamine. Mental illness was reported in 64% of adults with opioid use disorder, and 24.5% had both a mental illness and a substance use disorder (Jones & McCance-Katz, 2019). The relationship between mental health, drug use and dependence is complex. In some instances, underlying mental health conditions contribute to use and dependence. In other contexts, the use of opioids, particularly where illicit opioid use is criminalised, contributes to negative mental health and physical consequences for people who use opioids (UNODC, 2019). Mental health and substance use are augmented by entry into and release from prison (UNODC, 2019). Management of opioid dependence should be expanded to provide comprehensive care that addresses both opioid use and mental health (Jones & McCance-Katz, 2019).

Persons with opioid-related disorders lost almost eight life years before age 85 compared to persons without opioid-related disorders. The increase in mortality among opioid users compared to the general population observed is reported worldwide (Bahji A, Cheng B, Gray S, 2020; Larney et al., 2020). In high-income countries, excess mortality in persons with opioid dependence is driven by overdose, suicide, violence, and infectious disease complications (Bahji A, Cheng B, Gray S, 2020; Larney et al., 2020). However, in our study setting, the causes of mortality are different. The vast majority of people with opioid-related disorders died from natural causes and excess mortality due to unnatural causes contributed a negligible amount to the total excess mortality. In the South African context, opioids are more commonly smoked than injected, and there is limited access to illicit pharmaceutical opioids. These factors likely reduce the risk of overdose.

The total amount of methadone prescribed to beneficiaries suggests that dosing was likely lower than the recommended therapeutic doses and few, if any, received OAMT. The high cost of methadone has been cited as one reason limiting access in South Africa (Scheibe et al., 2018). This finding raises concerns about the management practices for opioid dependence in South Africa’s private sector. The coverage of the high cost of methadone by medical insurance schemes is very limited. Due to the high cost of opioid agonist medication, and the need for at least monthly doctor visits, it is likely that few clients can afford OAMT at the recommended dosages and duration.

The treatment of opioid dependence with OAMT is supported by strong evidence globally and in South Africa (Gloeck et al., 2020; Marks et al., 2020; Scheibe, Shelly, Gerardy, et al., 2020; Lize Weich, 2010; WHO, 2004). Long term use of OAMT (≥ 6 months) is associated with reduced dependence, reduced morbidity, and mortality from infectious diseases, reduced criminal behaviour, improved physical and psychological health and better social integration and functioning (Gloeck et al., 2020). Scaling up of harm reduction programmes that include OAMT is feasible in South Africa (Scheibe et al., 2018). Key areas to advance these goals would comprise; the inclusion of opioid agonists for use as OAMT on South Africa’s primary care level essential medicines list, the reduction of the cost of opioid agonist medication, open dissemination of research and service data on interventions for managing opioid dependence, and decriminalising people who use illicit substances (Scheibe et al., 2017, 2018).

The implications of our research include the need for healthcare providers to be capacitated to recognise, diagnose, and manage opioid-related disorders, guidelines and policies that ensure consistent care and adequate funding from medical insurance schemes for OAMT, and more funding and research to continue studying risk factors, treatment, health outcomes and prevention measures for opioid use.

This study contributes data on the management of opioid-related disorders from a large national private sector cohort. This is an important strength because data from the private sector on the management of opioid-related disorders is largely missing in South Africa. The results include data from all levels of care including outpatient care, in-hospital care, and pharmacy claims data. Linkage to the NPR also enables reliable ascertainment of mortality (Johnson et al., 2015). Improving the estimates of opioid-related disorders is essential for South Africa to respond to increasing level of opioid use.

The results should, however, be interpreted in light of the following limitations. First, our study likely underestimates the incidence of opioid-related disorders because our case definition relied on diagnoses and treatment as proxies for opioid-related disorders. In South Africa, mental health and substance use disorders often remain undiagnosed and untreated (Ruffieux et al., 2021), and our definition of opioid-related disorders would miss individuals with undiagnosed and untreated opioid-related disorders. The stigma attached to substance use disorders (P.W. Corrigan & Bink, 2016; Patrick W. Corrigan & Nieweglowski, 2018) and health care providers’ limited knowledge of the diagnostic criteria for opioid-related disorders (Daniels et al., 2021) may contribute to the underdiagnoses of opioid-related disorders. Second, we may have misclassified persons with opioid-related disorders if opioid agonists were used for indications other than treatment of opioid dependence or withdrawal or if ICD-10 codes were miscoded. We minimised misclassification from opioid agonist prescription by limiting the opioid agonists considered for identifying persons with opioid-related disorders to those indicated primarily for the treatment of opioid dependence and withdrawal management and not as pain medication. Third, our study only included data from a private-sector medical insurance scheme; thus, our findings do not necessarily apply to the public sector.

## Conclusions

The incidence of opioid-related disorders is increasing in South Africa’s private sector healthcare population, particularly among men and young adults and is associated with substantial psychiatric morbidity and excess mortality. The implementation of evidence-based management guidelines for opioid-related disorders is urgently needed to improve the care and health outcomes of people with opioid-related disorders.

## Data Availability

All data were obtained from the IeDEA-SA. Data cannot be made available online because of legal and ethical restrictions. To request data, readers may contact IeDEA-SA for consideration by filling out the online form available at https://www.iedea-sa.org/contact-us/.

## Author Contributions

MT, AS, and AH conceived the study and wrote the first draft of the study protocol. All authors contributed to the final version of the protocol. AH, YR, and MT performed statistical analysis. MT and AH wrote the first draft of the manuscript, which was revised by all authors. All authors approved the final version of the paper for submission.

## Ethical Standards

The authors assert that all procedures contributing to this work comply with the ethical standards of the relevant national and institutional committees on human experimentation and with the Helsinki Declaration of 1975, as revised in 2000.

---

**Figure S1.**
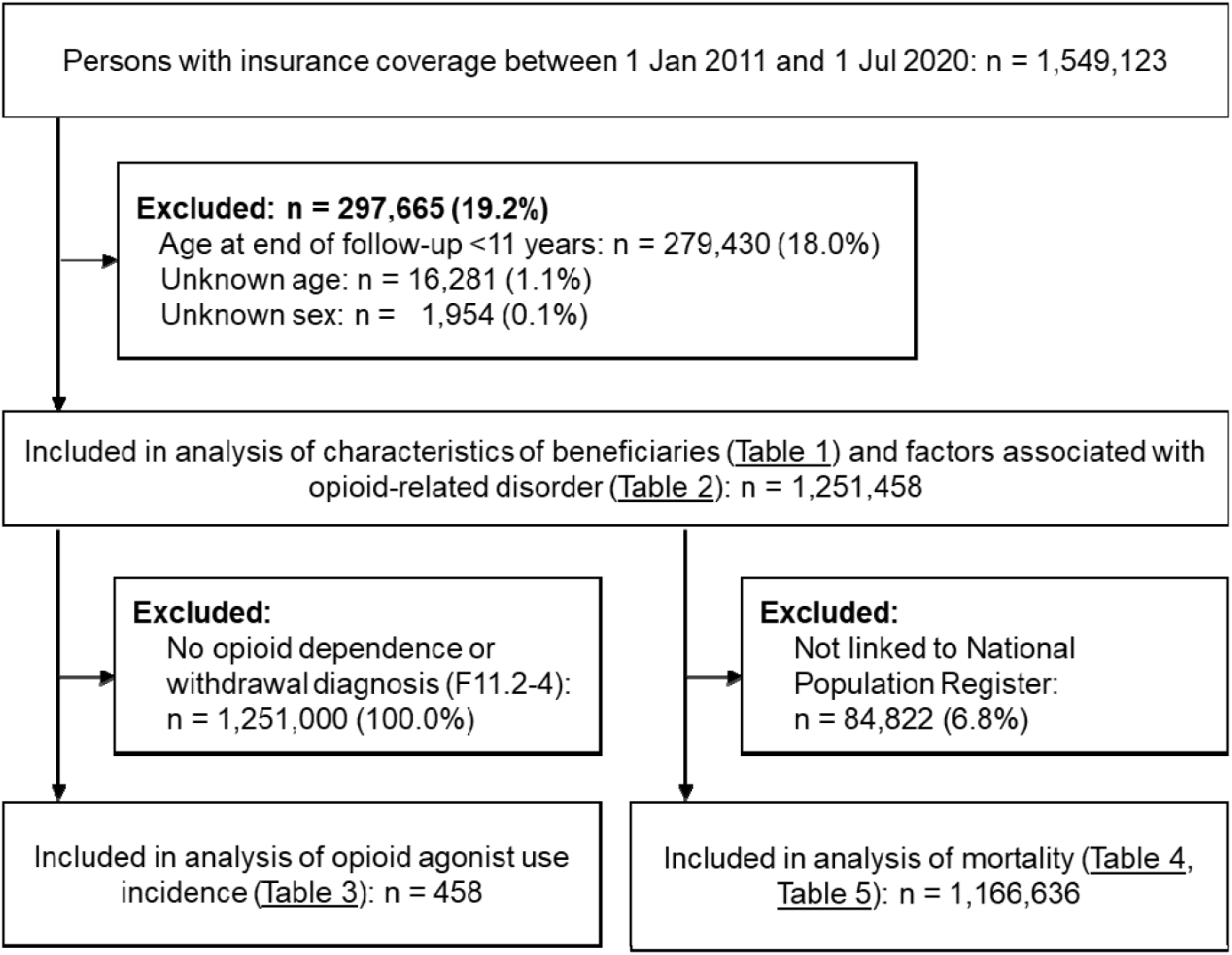
Flowchart of inclusion of beneficiaries in analyses.

